# Biomarkers of Length of Stay on an Inpatient Eating Disorder Unit

**DOI:** 10.1101/2020.12.17.20248429

**Authors:** Courtney E. Breiner, Baiyu Qi, Laura M. Thornton, Kimberly A. Brownley, Tonya Foreman, Anna M. Bardone-Cone, Cynthia M. Bulik, Jessica H. Baker

## Abstract

Length of stay on an inpatient unit for treatment of anorexia nervosa (AN) is widely variable. Although previous research has used anthropometric and clinical variables and duration of illness to predict length of stay, there has been limited investigation of the predictive ability of biomarkers. Biomarkers, including those collected through a comprehensive metabolic panel (CMP) and appetite hormones, such as ghrelin and leptin, are impacted by disease presence and may play an etiological role in AN. Using a series of regression models, we evaluated the associations of these putative biomarkers with length of inpatient stay in 46 females receiving treatment on an inpatient eating disorder unit. Active ghrelin levels at inpatient admission positively predicted length of stay and alkaline phosphatase at discharge was significantly positively correlated with length of stay. This research provides further evidence supporting both biological and psychological components of AN, identifying potential biomarkers that could aid in prospective prediction of treatment needs. Further research is necessary to replicate and extend these findings across treatment settings.

## 1. Introduction

Anorexia nervosa (AN) has one of the highest mortality rates of any mental illness and is more deadly than other eating disorders (Smink et al, 2012; Fichter & Quadflieg, 2016). AN involves severe psychological and medical sequelae (Westmoreland et al., 2016), and consequences of AN, such as extreme underweight status, malnutrition, nutrient deficiencies, and other medical complications, often require inpatient treatment for medical stabilization and psychological treatment. Directly conflicting with insurance-driven institutional efforts to shorten inpatient stays to decrease treatment costs (Wiseman et al., 2001; Weissman & Rosselli, 2017), premature discharge from inpatient treatment for AN leads to patients discharging at lower weights than may be medically recommended (Vandereycken, 2003; Sly et al., 2013) and to adverse outcomes, such as requiring additional hospitalization(s) after discharge (Pike, 1998). Together, this may ultimately lead to higher rates of inpatient readmission and, paradoxically, additional treatment costs (Vandereycken, 2003). Reliable, prospective predictors of inpatient length of stay could improve inpatient clinical management, yet few established predictors exist. Biomarkers may be particularly useful for predicting length of stay due to their objective nature and established associations with AN-illness status in combination with the fact that many biological indices are already tracked as part of routine clinical care in inpatient treatment settings. Thus, the primary goal of this study was to identify potential biomarkers of length of stay for patients with AN on an inpatient eating disorder unit.

Significant genetic correlations have been observed between AN and body mass index (BMI), insulin resistance, fasting insulin, leptin, high density lipoprotein (HDL) cholesterol, and type 2 diabetes suggesting that some of the genetic factors that influence AN and certain metabolic and arthrometric phenotypes are shared (Watson et al., 2019). Additionally, AN is associated with alterations in several metabolic indices. For example, patients with AN have increased insulin sensitivity compared with women without an eating disorder, which persists into the weight-restoration period (Dostálová et al., 2007; Brown et al., 2003; Kim et al., 2019; Ilyas et al., 2019).

Appetitive hormone dysregulation is also observed in AN (Hebebrand et al., 2007). Patients with AN have significantly *lower* leptin and *higher* active and total ghrelin levels than healthy controls (Schalla & Stengel, 2018). Leptin levels increase with weight gain in AN (Hebebrand et al., 2007; Ruscica et al., 2016), giving possible insight regarding whether a patient is recovering (i.e., together, weight and leptin levels are two indicators of treatment progress; Misra & Klibanski, 2010). Conversely, both active and total ghrelin levels decrease with weight gain after refeeding without psychological intervention (Schalla & Stengel, 2018; Nakahara et al., 2007). The genetic and phenotypic associations observed between multiple biological indices and AN suggest potential avenues for the identification of inpatient length of stay biomarkers. Examining the utility of biomarkers in predicting treatment course is a natural extension of this research. It is important to note that although these biomarkers may be related to starvation rather than AN pathophysiology per se, they remain important to examine as low weight is often a primary reason for inpatient treatment.

The goal of this study was to examine whether peripheral biomarkers from blood samples of AN inpatient length of stay could be identified. Peripheral biomarkers from blood have the benefit of being able to index biological factors from a variety of systems (e.g., stomach, kidneys, endocrine system). Here, we included biological indices typically captured as part of a comprehensive metabolic panel (CMP)—given this is often routinely monitored during inpatient treatment—and the appetite hormones ghrelin and leptin. The only *a priori* hypotheses we had were in regard to appetite hormones: namely that lower levels of leptin and higher levels of ghrelin at admission and discharge would be significantly associated with a longer length of stay. We considered the inclusion of the CMP as exploratory. Identifying biomarkers of inpatient length of stay for AN may provide additional information for AN treatment monitoring and stabilization.

## 2. Methods

### 2.1. Study population

Patients were recruited from an inpatient eating disorder unit at an academic medical center. Participants (N=73) were females at least 15 years of age who were diagnosed with AN by the inpatient unit psychiatrist using standard semi-structured interviewing procedures. One patient withdrew consent. Of the 72 remaining patients, a total of 46 participants who had blood drawn within five days after admission (T1) and at inpatient discharge (T2) and had complete data for the biomarkers of interest were included in the current analyses. This study was approved by the Biomedical Institutional Review Board at the University of North Carolina at Chapel Hill. Along with providing informed consent to participate in the study, participants also signed a HIPAA consent form so that treatment information (e.g., chemistry labs, admission height/weight, discharge date) could be obtained from the medical records.

### 2.2. Patient characteristics

Age at admission, height, and weight were obtained for all participants. Inpatient staff measured height and weight at admission and discharge. BMI (kg/m^2^) was calculated based on these values. Ideal body weight (IBW) was calculated as [100+5*(height in inches - 60)]; %IBW was calculated as (weight / IBW) *100.

### 2.3. Length of stay

Length of stay, the primary outcome variable, was defined as the number of days a participant spent on the inpatient unit. This was calculated as discharge date minus admission date. Premature discharge status was also coded: discharged by the treatment team or discharged against medical advice [AMA], which was considered premature/early by the treatment team.

### 2.4. Blood samples

Blood samples were measured at admission and prior to discharge. Fasting blood draws occurred in the morning, prior to breakfast. Blood samples for the CMP, as well as magnesium and phosphorus, were delivered to and processed by UNC Healthcare McClendon Hospital Laboratory Services (CLIA-certified). The CMP included the following: sodium, potassium, chloride, bicarbonate, urea nitrogen, creatinine, glucose, calcium, total protein, albumin, total bilirubin, aspartate aminotransferase, alanine aminotransferase, and alkaline phosphatase levels.

Additional blood samples were collected for leptin, total and active ghrelin, and estradiol assays at T1 and T2. Blood samples for hormone assays were obtained during regularly scheduled blood draws at T1 and T2. Total ghrelin (pg/ml), active ghrelin (pg/ml), and leptin (ng/ml) plasma hormone levels were measured using double antibody Radioimmunoassay (RIA) reagents and protocols commercially available from EMD Millipore, Billerica, MA. Intra-assay coefficients of variability (CV’s) are 7.9%, 6.7%, and 4.6% respectively, and inter-assay CV’s are 14.7%, 9.6%, and 5.0%. The sensitivity of the total ghrelin assay is 93 pg/ml with a standard range of 120-7700 pg/ml. Active ghrelin assay has a sensitivity of 7.8 pg/ml with a standard range of 6.9-1770 pg/ml. The sensitivity of the leptin RIA is .437ng/ml, and the standard range is .78 to 100 ng/ml. Plasma 17B-estradiol levels were measured using a coated tube RIA kit from MPBiomedicals, Costa Mesa, CA. The intra- and inter-assay CV’s are 3.5% and 7.6% respectively, and the sensitivity is 4 pg/ml with a standard range of 10-3000 pg/ml.

Given our interest in translatability to clinical management, we compared the value of potential biomarkers of length of stay to standard clinical reference ranges. UNC Healthcare McClendon Laboratory provided references ranges for the CMP values. For any lab parameter that provided age-specific reference ranges, adult ranges were used to interpret sample means given the varied age of the sample. Although empirical reference ranges have not often been used in appetite hormone research, clinical reference ranges do exist. Fasting reference ranges for total ghrelin and range for leptin were obtained from Mayo Clinic Laboratories, where enzyme immunoassay (EIA)/enzyme-linked immunosorbent assay (ELISA) was used for testing total ghrelin, and ELISA was used for testing leptin. Range for estradiol was obtained from UNC Healthcare McClendon Laboratory, where EIA was used. Notably, there are high correlations between total ghrelin measurements obtained with RIA and EIA (r=0.99; EMD Millipore), and between leptin measurements obtained with RIA and ELISA (r=0.96; Carlson et al., 1999). The concordance correlation coefficient showed a fair agreement between RIA and EIA for estradiol measurement (0.68, Skenandore et al., 2017). Therefore, these reference ranges are likely comparable to the current study.

### 2.5. Statistical analysis

Statistical analyses were conducted in SAS 9.4 (SAS Inc.). Descriptive information was generated for patient characteristics and potential biomarkers at both admission and discharge. Differences between T1 and T2 for each of these variables were evaluated using paired t-tests. We applied generalized linear models, using PROC GLM, to predict length of stay on the inpatient unit from each biomarker at T1 and T2, respectively. Admission age and BMI were included as covariates in each model.

Based on the results of the T1 and T2 models independently, we next used backward stepwise regression to obtain a more parsimonious prediction model, and to understand the importance of each biomarker in determining length of stay. This model included all significant biomarkers (*p*<0.05) from the linear regression models for both T1 and T2. Using stepwise regression allowed us to identify the best set of biomarkers to predict length of stay that we might evaluate further for clinical use. Age and BMI at admission were also included in the model as covariates but were allowed to leave the model in the elimination process. The variable with the least significant effect that did not meet the 0.05 significance threshold for staying in the model was removed from the model. The elimination process was repeated until no improvement of the model was achieved by removing new variables, and all remaining effects in the model met the significance threshold (*p*<0.05) (Heinze et al., 2018). Adjusted R^2^ were calculated to measure the proportion of the variance for length of stay that was explained by the final model. Standardized regression coefficients were calculated to estimate and compare the individual effects of each predictor.

Finally, based on the number of AMA discharges observed, a post-hoc sensitivity analysis was conducted to assess the impact of these patients on the results. Specifically, generalized linear models and a backward stepwise regression model were conducted for patients without premature discharge only to evaluate whether the same biomarkers that predicted length of stay in the full sample remained significant for patients who did not discharge AMA. Due to our limited sample size, in addition to statistical significance, we also used standardized regression coefficients to measure effect sizes.

Notably, because we considered this study exploratory, we decided *a priori* to not correct for multiple comparisons. Correction for multiple comparisons increases the risk for type-2 error (i.e., non-rejection of a false null hypothesis), which may prematurely discard useful observations (Rothman, 1990). Here, we judged potential clinical utility as more important than statistical significance. Future studies will need to confirm any observed associations.

## 3. Results

### 3.1. Demographics

**Table 1** summarizes patient characteristics at admission and discharge. Our study sample consisted of 19 (41.30%) patients with restricting-type AN, 25 (54.35%) patients with binge-eating/purging-type AN, and 2 (4.35%) patients with atypical AN. The average age of the patients was 29.63 (*SD*=14.20) years. BMI increased significantly from admission (14.93 kg/m^2^) to discharge (17.09 kg/m^2^). At discharge, patients had a mean %IBW of 82.72 (*SD*=7.59); only 18 (39.13%) patients achieved ≥ 85% IBW. The mean length of stay on the inpatient unit was 33.52 (*SD*=18.07) days. Thirteen (28.26%) patients discharged AMA. Of those patients who discharged early, the mean length of inpatient stay was 32.92 (19.39) days, with a range of 10 to 78 days.

**Table 1.**
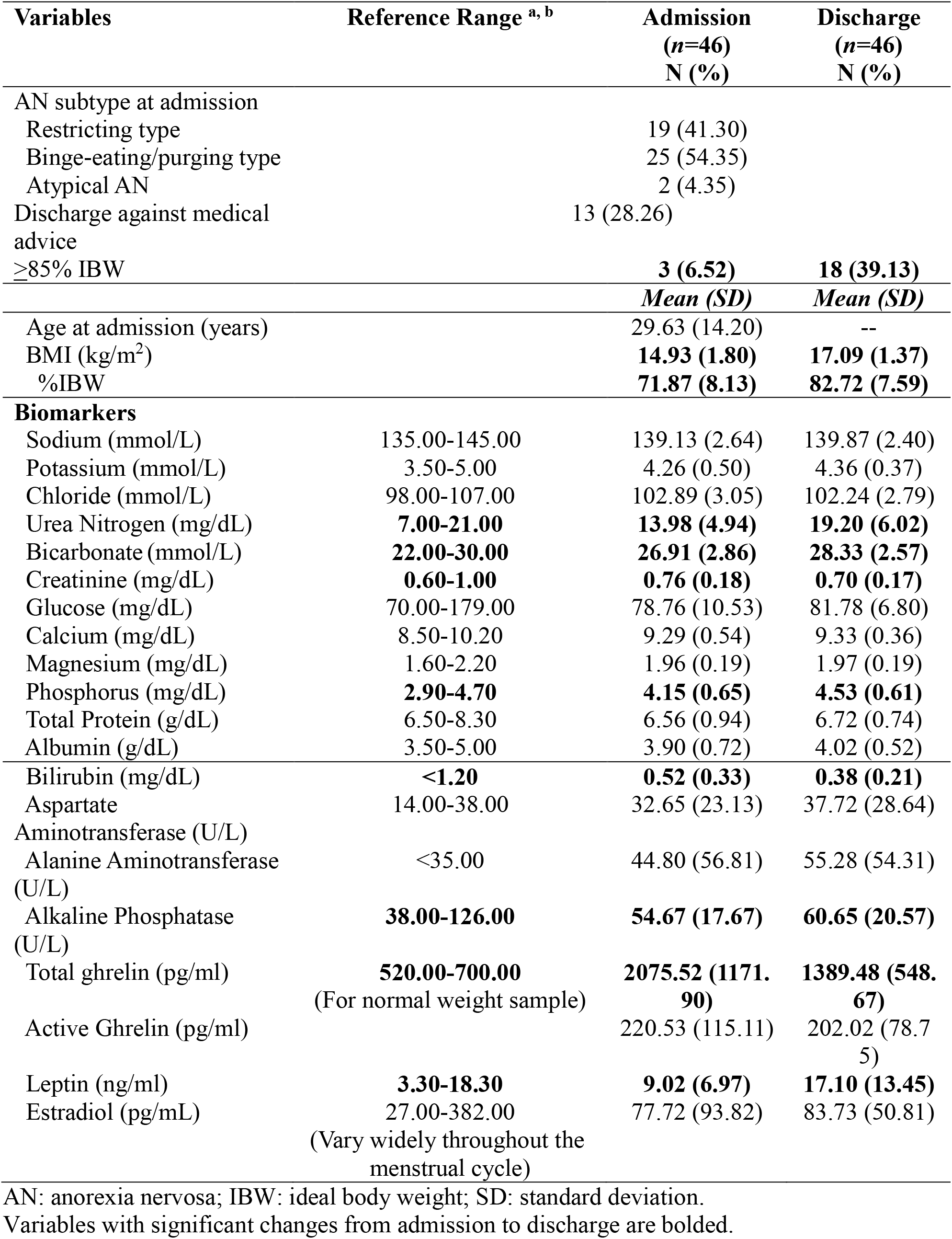
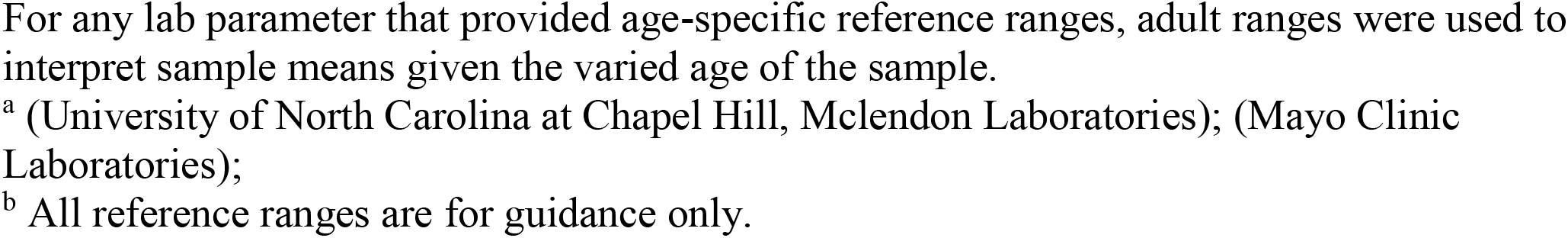
Patient Characteristics at Inpatient Admission and Discharge.

| Variables | Reference Range <sup>a, b</sup> | Admission<br>(*n*=46)<br>N (%) | Discharge<br>(*n*=46)<br>N (%) |
| --- | --- | --- | --- |
| AN subtype at admission |  |  |  |
| Restricting type |  | 19 (41.30) |  |
| Binge-eating/purging type |  | 25 (54.35) |  |
| Atypical AN |  | 2 (4.35) |  |
| Discharge against medical advice |  | 13 (28.26) |  |
| ≥85% IBW |  | **3 (6.52)** | **18 (39.13)** |
|  |  | *Mean (SD)* | *Mean (SD)* |
| Age at admission (years) |  | 29.63 (14.20) | -- |
| BMI (kg/m<sup>2</sup>) |  | **14.93 (1.80)** | **17.09 (1.37)** |
| %IBW |  | **71.87 (8.13)** | **82.72 (7.59)** |
| **Biomarkers** |  |  |  |
| Sodium (mmol/L) | 135.00-145.00 | 139.13 (2.64) | 139.87 (2.40) |
| Potassium (mmol/L) | 3.50-5.00 | 4.26 (0.50) | 4.36 (0.37) |
| Chloride (mmol/L) | 98.00-107.00 | 102.89 (3.05) | 102.24 (2.79) |
| Urea Nitrogen (mg/dL) | **7.00-21.00** | **13.98 (4.94)** | **19.20 (6.02)** |
| Bicarbonate (mmol/L) | **22.00-30.00** | **26.91 (2.86)** | **28.33 (2.57)** |
| Creatinine (mg/dL) | **0.60-1.00** | **0.76 (0.18)** | **0.70 (0.17)** |
| Glucose (mg/dL) | 70.00-179.00 | 78.76 (10.53) | 81.78 (6.80) |
| Calcium (mg/dL) | 8.50-10.20 | 9.29 (0.54) | 9.33 (0.36) |
| Magnesium (mg/dL) | 1.60-2.20 | 1.96 (0.19) | 1.97 (0.19) |
| Phosphorus (mg/dL) | **2.90-4.70** | **4.15 (0.65)** | **4.53 (0.61)** |
| Total Protein (g/dL) | 6.50-8.30 | 6.56 (0.94) | 6.72 (0.74) |
| Albumin (g/dL) | 3.50-5.00 | 3.90 (0.72) | 4.02 (0.52) |
| Bilirubin (mg/dL) | **<1.20** | **0.52 (0.33)** | **0.38 (0.21)** |
| Aspartate | 14.00-38.00 | 32.65 (23.13) | 37.72 (28.64) |
| Aminotransferase (U/L) |  |  |  |
| Alanine Aminotransferase (U/L) | <35.00 | 44.80 (56.81) | 55.28 (54.31) |
| Alkaline Phosphatase (U/L) | **38.00-126.00** | **54.67 (17.67)** | **60.65 (20.57)** |
| Total ghrelin (pg/ml) | **520.00-700.00** | **2075.52 (1171.90)** | **1389.48 (548.67)** |
|  | (For normal weight sample) |  |  |
| Active Ghrelin (pg/ml) |  | 220.53 (115.11) | 202.02 (78.75) |
| Leptin (ng/ml) | **3.30-18.30** | **9.02 (6.97)** | **17.10 (13.45)** |
| Estradiol (pg/mL) | 27.00-382.00 | 77.72 (93.82) | 83.73 (50.81) |
|  | (Vary widely throughout the menstrual cycle) |  |  |

AN: anorexia nervosa; IBW: ideal body weight; SD: standard deviation.
Variables with significant changes from admission to discharge are bolded.
For any lab parameter that provided age-specific reference ranges, adult ranges were used to interpret sample means given the varied age of the sample.
<sup>b</sup> All reference ranges are for guidance only.

Compared with admission, patients at discharge had statistically significantly *lower* mean levels of creatinine and bilirubin, and statistically significantly *higher* mean levels of urea nitrogen, bicarbonate, phosphorus, alkaline phosphatase, and leptin. However, values for these measures were within the clinically acceptable ranges at both time points (University of North Carolina at Chapel Hill, McLendon Laboratories; Mayo Clinic Laboratories). Of note, patients’ mean alanine aminotransferase levels were above the clinically acceptable range at both time points, with no significant change over time. The mean level of total ghrelin was also above the clinical range at both time points, and there was a statistically significant decrease from T1 to T2.

### 3.2. Linear regression results

**Table 2** shows the results from the generalized linear regression models predicting length of stay on the inpatient unit from each biomarker at admission and discharge, respectively. At T1, *lower* magnesium and *higher* active ghrelin were significantly associated with a longer length of stay on the inpatient unit, after adjusting for admission age and BMI. At T2, *higher* glucose, alkaline phosphatase, and leptin were significantly associated with a longer length of stay on the inpatient unit, after adjusting for age and admission BMI.

**Table 2.** Results from generalized linear regression analyses predicting length of stay on the inpatient unit from each biomarker at admission and discharge, adjusting for age and BMI at admission

| **Variable** | **DF** | **B** | **SE** | **$\beta$** | **t value** | **p-value** |
| --- | --- | --- | --- | --- | --- | --- |
| Biomarkers at admission |  |  |  |  |  |  |
| Sodium | 1 | 1.70 | 0.81 | 0.17 | 1.44 | 0.16 |
| Potassium | 1 | 0.76 | 4.19 | 0.02 | 0.18 | 0.86 |
| Chloride | 1 | 0.10 | 0.65 | 0.02 | 0.15 | 0.88 |
| Urea Nitrogen | 1 | 0.32 | 0.40 | 0.09 | 0.79 | 0.44 |
| Bicarbonate | 1 | 0.22 | 0.69 | 0.03 | 0.31 | 0.76 |
| Creatinine | 1 | -8.77 | 10.81 | -0.09 | -0.81 | 0.42 |
| Glucose | 1 | -0.02 | 0.21 | -0.01 | -0.08 | 0.94 |
| Calcium | 1 | 5.41 | 4.37 | 0.16 | 1.24 | 0.22 |
| **Magnesium** | 1 | -22.33 | 9.90 | -0.23 | -2.26 | 0.03 |
| Phosphorus | 1 | -2.23 | 3.11 | -0.08 | -0.72 | 0.48 |
| Total Protein | 1 | 0.56 | 2.41 | 0.03 | 0.23 | 0.82 |
| Albumin | 1 | 0.65 | 3.18 | 0.03 | 0.21 | 0.84 |
| Bilirubin | 1 | 8.31 | 5.94 | 0.15 | 1.40 | 0.17 |
| Aspartate | 1 | 0.06 | 0.09 | 0.08 | 0.74 | 0.47 |
| Aminotransferase |  |  |  |  |  |  |
| Alanine | 1 | 0.04 | 0.04 | 0.14 | 1.21 | 0.23 |
| Aminotransferase |  |  |  |  |  |  |
| Alkaline Phosphatase | 1 | 0.13 | 0.11 | 0.12 | 1.14 | 0.26 |
| Total Ghrelin | 1 | 0.003 | 0.001 | 0.22 | 1.76 | 0.09 |
| **Active Ghrelin** | 1 | 0.04 | 0.02 | 0.28 | 2.45 | 0.02 |
| Leptin | 1 | 0.18 | 0.29 | 0.07 | 0.63 | 0.53 |
| Estradiol | 1 | -0.004 | 0.02 | -0.02 | -0.20 | 0.84 |
| Biomarkers at discharge |  |  |  |  |  |  |
| Sodium | 1 | 0.11 | 0.84 | 0.02 | 0.13 | 0.89 |
| Potassium | 1 | -8.85 | 5.42 | -0.18 | -1.63 | 0.11 |
| Chloride | 1 | -0.77 | 0.71 | -0.12 | -1.08 | 0.29 |
| Urea Nitrogen | 1 | 0.07 | 0.44 | 0.02 | 0.17 | 0.87 |
| Bicarbonate | 1 | 1.92 | 0.76 | 0.13 | 1.21 | 0.23 |
| Creatinine | 1 | -14.97 | 12.66 | -0.14 | -1.18 | 0.24 |
| **Glucose** | 1 | 0.79 | 0.26 | 0.30 | 2.99 | 0.01 |
| Calcium | 1 | 6.60 | 5.75 | 0.13 | 1.15 | 0.26 |
| Magnesium | 1 | -16.50 | 10.23 | -0.17 | -1.61 | 0.11 |
| Phosphorus | 1 | -0.52 | 3.31 | -0.02 | -0.16 | 0.88 |
| Total Protein | 1 | 4.93 | 2.75 | 0.20 | 1.79 | 0.08 |
| Albumin | 1 | 7.19 | 4.07 | 0.21 | 1.77 | 0.09 |
| Bilirubin | 1 | 15.04 | 10.24 | 0.17 | 1.47 | 0.15 |
| Aspartate | 1 | 0.07 | 0.07 | 0.11 | 1.03 | 0.31 |
| Aminotransferase |  |  |  |  |  |  |
| Alanine | 1 | 0.04 | 0.04 | 0.11 | 1.05 | 0.30 |
| Aminotransferase |  |  |  |  |  |  |
| **Alkaline Phosphatase** | 1 | 0.32 | 0.09 | 0.36 | 3.66 | <0.01 |
| Total Ghrelin | 1 | -0.002 | 0.004 | -0.06 | -0.52 | 0.60 |
| Active Ghrelin | 1 | 0.04 | 0.02 | 0.10 | 1.67 | 0.10 |
| **Leptin** | 1 | 0.31 | 0.14 | 0.23 | 2.26 | 0.03 |
| Estradiol | 1 | 0.03 | 0.04 | 0.08 | 0.78 | 0.44 |

DF: degree of freedom; B: regression coefficient; SE: standard error; $\beta$ : standardized regression coefficient;
Variables with significant association ( $p < 0.05$ ) with length of stay are bolded.

### 3.3. Stepwise regression results

Backward stepwise regression was used to identify which biomarkers played the most significant roles in determining length of stay on the inpatient unit (**Table 3**). Significant biomarkers from generalized linear regression models for T1 (magnesium and active ghrelin) and T2 (glucose, alkaline phosphatase, and leptin) were entered into the model along with age and BMI at admission. Active ghrelin at T1, alkaline phosphatase at T2, and BMI at T1 were retained in the final model and jointly accounted for 65% of the variance in length of stay. BMI at T1 (*β*=-0.53) had the largest effect on length of stay, followed by alkaline phosphatase at T2 (*β*=0.35) and active ghrelin at T1 (*β*=0.26).

**Table 3.** Biomarkers retained in the backward stepwise regression model predicting length of stay on the inpatient unit

| **Parameter** | ***DF*** | ***B*** | ***SE*** | **$\beta$** | ***t value*** | ***p-value*** | **Adj R-Sq** |
| --- | --- | --- | --- | --- | --- | --- | --- |
| Active Ghrelin at admission | 1 | 0.04 | 0.01 | 0.26 | 2.80 | 0.01 | 0.65 |
| Alkaline Phosphatase at discharge | 1 | 0.30 | 0.08 | 0.35 | 3.73 | <0.01 |  |
| BMI at admission | 1 | -5.31 | 0.95 | -0.53 | -5.62 | <0.01 |  |

DF: degree of freedom; B: regression coefficient; SE: standard error; $\beta$ : standardized regression coefficient; Adj R-Sq: adjusted r-squared

### 3.4. Post-hoc sensitivity analysis

Given the number of participants who terminated treatment early (*n*=13), a sensitivity analysis was conducted to address the impact of these individuals on the results. Specifically, analyses were repeated including only patients with a treatment team approved discharge (*n*=33) (**Table 4)**. We did not identify any new significant biomarkers of length of stay in this subgroup. At T1, magnesium (*β* changed from −0.23 to −0.17) and active ghrelin (*β* changed from 0.28 to 0.23) were no longer significantly associated with length of stay. At T2, *higher* glucose (*β* changed from 0.30 to 0.32) and alkaline phosphatase (*β* changed from 0.36 to 0.28) remained significantly associated with longer length of stay. However, leptin (*β* changed from 0.23 to 0.25) was no longer significantly associated with length of stay. For the final stepwise model, BMI at T1 (*β*=-0.70) and alkaline phosphatase at T2 (*β*=0.35) were retained, jointly accounting for 55% of the variance in length of stay. Taken together, some biomarkers of length of stay that were identified in the total sample (magnesium at T1, active ghrelin at T1, and leptin at T2) were no longer significant in the sensitivity analysis. However, the effect sizes did not change much indicating that the loss of significance may be due to reduced statistical power.

**Table 4.**
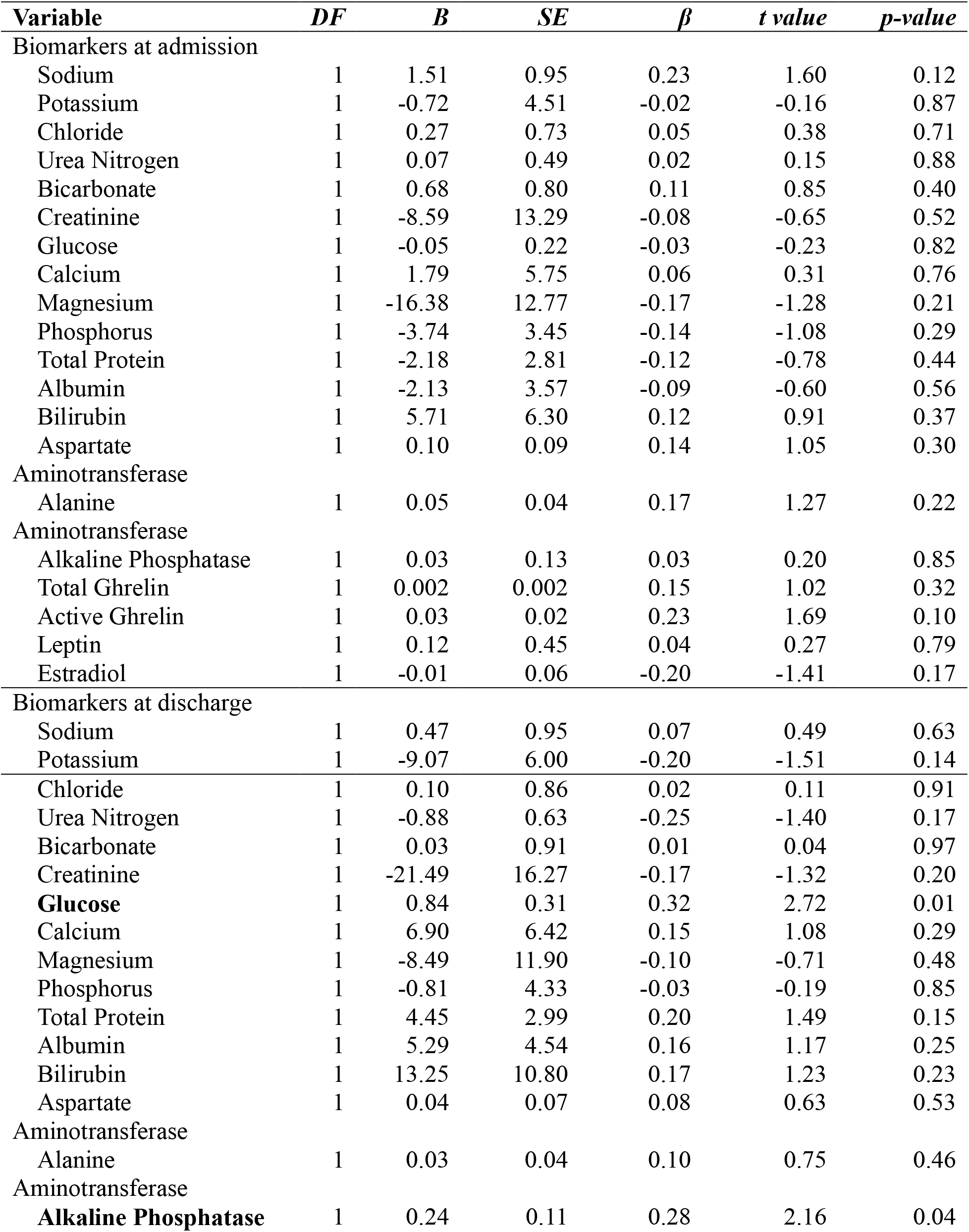
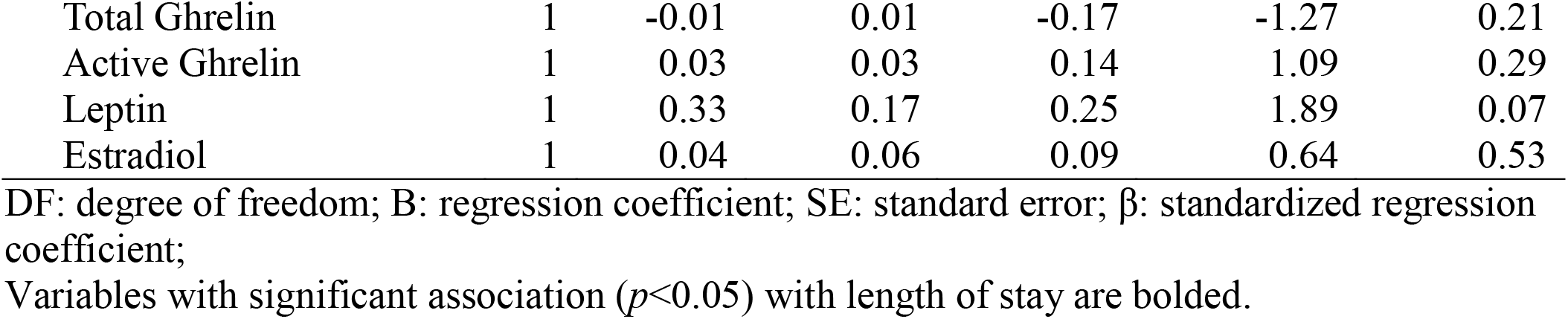
Post-hoc Sensitivity Analyses: Generalized linear regression results predicting length of stay on the inpatient unit from each biomarker at admission removing patients with premature discharge, adjusting for age and BMI at admission

## 4. Discussion

This study addresses an important gap in the literature, namely whether peripheral blood-based biomarkers associated with length of stay can be identified. In the initial analyses, two variables at admission predicted longer length of stay on an inpatient unit: lower magnesium and higher active ghrelin. At discharge, three variables, higher glucose, higher leptin, and higher alkaline phosphatase, were associated with longer length of stay. Including all variables in a single stepwise regression model identified higher active ghrelin at admission and higher alkaline phosphatase at discharge as significantly associated with a longer length of stay, along with admission BMI. Together, active ghrelin at admission and alkaline phosphatase at discharge accounted for 65% of the variance in length of stay. Our findings highlight the potential for using biological indices as additional indicators of length of stay and implicate metabolic traits as contributors to treatment course.

In particular, our results indicate the potential utility of active and total ghrelin monitoring, which is not routinely assessed during AN treatment. Active ghrelin represents the acetyl isoform of ghrelin and is an appetite stimulant, rising before food intake, along with involvement in a number of other physiological functions such as lipid storage, body weight, energy expenditure, learning and memory and mood and anxiety (Hillman et al., 2011; Müller et al., 2015). Active ghrelin has a short half-life and is subsequently converted to non-acetyl (i.e., inactive) ghrelin (Tong et al., 2013). Total ghrelin represents active plus inactive ghrelin. Together, our results are generally consistent with previous findings suggesting that patients with AN have increased plasma ghrelin levels compared with those without an eating disorder (Méquinion et al., 2013).

Indeed, we observed that higher *active* ghrelin levels at admission predicted longer treatment stays. Although total ghrelin was not retained in the final regression model predicting inpatient stay, we also observed a significant decrease in *total* ghrelin levels from admission to discharge. Despite this decrease, total ghrelin levels at both admission and discharge were higher than a provided reference range (Mayo Clinic Laboratories), suggesting that although there was a statistically significant change, there may not be clinically significant change in these levels. Our results are consistent with previous findings suggesting that patients who have completed only inpatient medical stabilization treatment for AN have significantly higher total ghrelin levels than healthy controls even after nutritional rehabilitation and achieving weight gain within 10% of their goal weight on an inpatient unit (Nakahara et al., 2007). Our results also replicate research showing that total ghrelin levels trend downwards at one-year follow-up from weight restoration (Misra et al., 2005). However, contrary to these results, Otto and colleagues (2001) found that total ghrelin levels did not normalize after an inpatient refeeding protocol.

Taken together, ghrelin results suggest that patients with persistent elevations in active ghrelin may need longer inpatient stays to ensure medical stabilization and appropriateness for discharge whereas nutritional rehabilitation and initial weight gain do not necessarily equate with metabolic “recovery” as indexed by total ghrelin. Because ghrelin is involved in a number of indices likely relevant to AN treatment (e.g., appetite, mood, weight), this could suggest that persistently elevated ghrelin levels (i.e., active and total) may represent a biomarker of more severe, chronic, or metabolically unstable AN symptomatology requiring longer inpatient stays. Although speculative at this point, this hypothesis is supported by the fact both active and total ghrelin levels were persistently elevated and active ghrelin was associated with a longer length of stay.

Lower alkaline phosphatase at inpatient discharge was also significantly associated with length of stay. Alkaline phosphatase is an homodimeric protein enzyme and is implicated in bone and liver health. Low levels of alkaline phosphatase are an indicator of liver functioning abnormalities and could indicate stunted anabolic processes (Nova et al., 2008), and low alkaline phosphatase has previously been implicated as a marker of past and current illness in AN (Milner et al., 1985; Umeki, 1988). Alkaline phosphatase is decreased in individuals with AN compared with healthy controls, can remain decreased for up to a year post-discharge from inpatient treatment (Nova et al., 2008), and is positively correlated with other outcome markers, such as BMI and percent body fat and is significantly higher in those who are in partial or complete recovery from AN compared with individuals who are not recovered (Legroux-Gerot et al., 2005). Despite these past findings, recent research has indicated that alkaline phosphatase remains within a normal range throughout treatment for AN (Narayanan et al., 2010; Rosen et al., 2016; Gibson et al., 2020). These results indicate potential discrepancies in how “normal” levels are defined, and whether or not control groups are used for comparison.

Collectively, our results indicate that biomarkers may be useful for predicting AN inpatient treatment length of stay. Although other disease-specific markers have been used previously [e.g., duration of illness, (Lievers at al., 2009), BMI and previous hospital admissions (Maguire et al., 2003), early treatment response (Wales et al., 2016), and autonomous control over treatment (Thaler et al., 2016)], biomarkers have the benefit of being objective measures that do not rely on self-report or subjective clinical judgment. Additionally, peripheral blood-based biomarkers can be easily integrated into routine collection procedures, and often standardized interpretation procedures are in place (e.g., clinical references ranges). In particular, our findings suggest active ghrelin could be a useful addition for regular monitoring throughout routine clinical care given the prospective nature of active ghrelin at admission ultimately predicting inpatient length of stay. Definitions of AN outcome and recovery may need to be reconceptualized to include hormone regulation and metabolic recovery.

Our results also question whether current clinical reference ranges are able to capture subtle nuances in AN patients and whether disease-specific ranges could be useful for AN patients. Relying on community-based references may lead us to overlook clinically-relevant *changes* occurring in biological indices in patients with AN. Reference ranges from an otherwise healthy community population may not be adequately sensitive to detect the impact of AN-disease on metabolic levels, for example, in an AN population, regardless of whether the absolute levels are outside of established clinical norms. Current reference ranges may make it difficult to detect which patients require a longer length of stay without a comparison group of other AN patients. For example, we found statistically significant changes in seven (creatinine, bilirubin, urea nitrogen, bicarbonate, phosphorus, alkaline phosphatase, and leptin) lab indices over the course of treatment. However, these values remained in the normative range at both admission and discharge; it is unclear if these values are statistically significant but not clinically significant, or if they had clinically significant changes within the reference range. It will be important to continue to study the levels of these indices within AN samples across various disease stages as well as age- and sex-matched healthy controls.

There are limitations of this study that should be considered. First, we had a small sample size (*n*=46), which limited our power to identify statistically significant biomarkers of length of stay. Particularly, after removing patients who were prematurely discharged (AMA), we were unable to replicate some significant findings (magnesium at T1, active ghrelin at T1, and leptin at T2). However, the effect sizes did not change much, indicating that we lacked power. Therefore, it is important for future studies to identify potential biomarkers of length of stay using a larger sample size. Second, we had 13 patients (28.26%) terminate treatment AMA: this is likely a fairly heterogenous group as the range of days spent on the inpatient unit was wide (10 to 78 days), indicating that this was not simply a group of individuals initiating discharge soon after admission. We do not have the specific details about the reasons for premature discharge (i.e., initiated by the patient or whether the stay was truncated by insurance coverage), as reasons for such discharge may modify observed effects (e.g., group interaction). Larger studies must be conducted to replicate these findings. Third, our study did not include a comparison of controls without a lived experience with eating disorders. Finally, our findings may not generalize across different levels of care, and a multi-site study including more frequent sampling and a longitudinal follow-up period would provide more robust results as we could evaluate differences across treatment teams, type of treatment (inpatient, residential, intensive outpatient, and outpatient), and locations.

Despite the limitations, the results of this study identified objective peripheral blood-based biomarkers associated with length of stay on an inpatient unit that may aid in identifying patients who require more time on an inpatient unit and with continued research, could lead to prospective identifiers of treatment needs and readiness for discharge. Particularly important, this study may be more representative of patients with AN given the research was conducted in a treatment setting and not in a strictly research setting (e.g., clinical trial) and thus, displays additional utility of biological indices already being collected throughout treatment. Finally, using these biomarkers may aid in our ability to argue for a longer length of inpatient stays for patients with AN, given that one concern for Americans with eating disorders is limited insurance coverage for inpatient treatment (Escobar-Koch et al., 2010). Future research should examine the predictive nature of the identified biomarkers across follow-up periods, and whether or not they remain associated with treatment outcomes beyond discharge. Long-term, biomarkers represent another tool that could be used in the clinical management of AN and the determination for the appropriateness of inpatient discharge, ultimately improving the likelihood for positive outcomes and full recovery.

## Supporting information

Checklist

## Data Availability

Data will be made available upon reasonable request

## Acknowledgements

Funding: This work was supported by the National Institutes of Health [R01 MH095860; K01 MH106675; R01 MH120170; R01 MH119084; R01 MH118278; U01 MH109528], the Brain and Behavior Research Foundation, the Swedish Research Council (Vetenskapsrådet. [538-2013-8864]), and the Lundbeck Foundation [R276-2018-4581].

## References

Brown, N. W., Ward, A., Surwit, R., Tiller, J., Lightman, S., Treasure, J. L., & Campbell, I. C. (2003). Evidence for metabolic and endocrine abnormalities in subjects recovered from anorexia nervosa. Metabolism-Clinical and Experimental, 52(3), 296–302.

Carlson, M. G., Snead, W. L., Oeser, A. M., & Butler, M. G. (1999). Plasma leptin concentrations in lean and obese human subjects and Prader-Willi syndrome: Comparison of RIA and ELISA methods. Journal of Laboratory and Clinical Medicine, 133(1), 75–80.

Dostálová, I., Smitka, K., Papežová, H., Kvasničková, H., & Nedvídková, J. (2007). Increased insulin sensitivity in patients with anorexia nervosa: The role of adipocytokines. Physiological Research, 56(5), 587–594.

Escobar-Koch, T., Banker, J. D., Crow, S., Cullis, J., Ringwood, S., Smith, G. … & Schmidt, U. (2010). Service users’ views of eating disorder services: an international comparison. International Journal of Eating Disorders, 43(6), 549–559.

Fichter, M. M., & Quadflieg, N. (2016). Mortality in eating disorders-results of a large prospective clinical longitudinal study. International Journal of Eating Disorders, 49(4), 391–401.

Gibson, D., Watters, A., Cost, J., Mascolo, M., & Mehler, P. S. (2020). Extreme anorexia nervosa: medical findings, outcomes, and inferences from a retrospective cohort. Journal of Eating Disorders, 8(1), 1–10.

Hebebrand, J., Muller, T. D., Holtkamp, K., & Herpertz-Dahlmann, B. (2007). The role of leptin in anorexia nervosa: Clinical implications. Molecular Psychiatry, 12(1), 23–35.

Heinze, G., Wallisch, C., & Dunkler, D. (2018). Variable selection – A review and recommendations for the practicing statistician. Biometrical Journal, 60(3), 431–449.

Hillman, J. B., Tong, J., & Tschop, M. (2011). Ghrelin biology and its role in weight-related disorders. Discovery Medicine, 11(61), 521–528.

Ilyas, A., Hübel, C., Stahl, D., Stadler, M., Ismail, K., Breen, G. … & Kan, C. (2019). The metabolic underpinning of eating disorders: A systematic review and meta-analysis of insulin sensitivity. Molecular and Cellular Endocrinology, 497, 110307.

Kim, Y., Hildebrandt, T., & Mayer, L. E. (2019). Differential glucose metabolism in weight restored women with anorexia nervosa. Psychoneuroendocrinology, 110, 104404.

Legroux-Gerot, I., Vignau, J., Collier, F., & Cortet, B. (2005). Bone loss associated with anorexia nervosa. Joint Bone Spine, 72(6), 489–495.

Lievers, L. S., Curt, F., Wallier, J., Perdereau, F., Rein, Z., Jeammet, P., & Godart, N. (2009). Predictive factors of length of inpatient treatment in anorexia nervosa. European Child & Adolescent Psychiatry, 18(2), 75–84.

Maguire, S., Surgenor, L. J., Abraham, S., & Beumont, P. (2003). An international collaborative database: its use in predicting length of stay for inpatient treatment of anorexia nervosa. Australian and New Zealand Journal of Psychiatry, 37(6), 741–747.

Mayo Clinic Laboratories. Retrieved from https://www.mayocliniclabs.com/index.html.

University of North Carolina at Chapel Hill, Mclendon Laboratories. McLendon Clinical Laboratories. Retrieved from https://www.uncmedicalcenter.org/mclendon-clinical-laboratories/.

Méquinion, M., Langlet, F., Zgheib, S., Dickson, S., Dehouck, B., Chauveau, C., & Viltart, O. (2013). Ghrelin: Central and peripheral implications in anorexia nervosa. Frontiers in Endocrinology, 4, 15.

Milner, M. R., McAnarney, E. R., & Klish, W. J. (1985). Metabolic abnormalities in adolescent patients with anorexia nervosa. Journal of Adolescent Health Care, 6(3), 191–195.

Misra, M., Miller, K. K., Kuo, K., Griffin, K., Stewart, V., Hunter, E. …, & Klibanski, A. (2005). Secretory dynamics of ghrelin in adolescent girls with anorexia nervosa and healthy adolescents. American Journal of Physiology-Endocrinology and Metabolism, 289(2), E347–E356.

Misra, M., & Klibanski, A. (2010). Neuroendocrine consequences of anorexia nervosa in adolescents. Pediatric Neuroendocrinology, 17, 197–214.

Müller, T. D., Nogueiras, R., Andermann, M. L., Andrews, Z. B., Anker, S. D., Argente, J. …, & Casanueva, F. F. (2015). Ghrelin. Molecular Metabolism, 4(6), 437–460.

Nakahara, T., Kojima, S., Tanaka, M., Yasuhara, D., Harada, T., Sagiyama, K. I. …, & Naruo, T. (2007). Incomplete restoration of the secretion of ghrelin and PYY compared to insulin after food ingestion following weight gain in anorexia nervosa. Journal of Psychiatric Research, 41(10), 814–820.

Narayanan, V., Gaudiani, J. L., Harris, R. H., & Mehler, P. S. (2010). Liver function test abnormalities in anorexia nervosa—cause or effect. International Journal of Eating Disorders, 43(4), 378–381.

Nova, E., Lopez-Vidriero, I., Varela, P., Casas, J., & Marcos, A. (2008). Evolution of serum biochemical indicators in anorexia nervosa patients: A 1-year follow-up study. Journal of Human Nutrition and Dietetics, 21(1), 23–30.

Otto, B., Cuntz, U., Fruehauf, E. A., Wawarta, R., Folwaczny, C., Riepl, R. L. …, & Tschöp, M. (2001). Weight gain decreases elevated plasma ghrelin concentrations of patients with anorexia nervosa. European Journal of Endocrinology, 145(5), 669–673.

Pike, K. M. (1998). Long-term course of anorexia nervosa: Response, relapse, remission, and recovery. Clinical Psychology Review, 18(4), 447–475.

Rosen, E., Sabel, A. L., Brinton, J. T., Catanach, B., Gaudiani, J. L., & Mehler, P. S. (2016). Liver dysfunction in patients with severe anorexia nervosa. International Journal of Eating Disorders, 49(2), 151–158.

Rothman, K. J. (1990). No adjustments are needed for multiple comparisons. Epidemiology, 1(1), 43–46.

Ruscica, M., Macchi, C., Gandini, S., Morlotti, B., Erzegovesi, S., Bellodi, L., & Magni, P. (2016). Free and bound plasma leptin in anorexia nervosa patients during a refeeding program. Endocrine, 51(2), 380–383.

SAS Institute Inc. (2004). SAS/STAT® Software: Version 9. Cary, NC: SAS Institute, Inc.

Schalla, M. A., & Stengel, A. (2018). The role of ghrelin in anorexia nervosa. International Journal of Molecular Sciences, 19(7), 2117.

Skenandore, C. S., Pineda, A., Bahr, J. M., Newell-Fugate, A. E., & Cardoso, F. C. (2017). Evaluation of a commercially available radioimmunoassay and enzyme immunoassay for the analysis of progesterone and estradiol and the comparison of two extraction efficiency methods. Domestic Animal Endocrinology, 60, 61–66.

Smink, F. R., Van Hoeken, D., & Hoek, H. W. (2012). Epidemiology of eating disorders: Incidence, prevalence and mortality rates. Current Psychiatry Reports, 14(4), 406–414.

Sly, R., Morgan, J. F., Mountford, V. A., & Lacey, J. H. (2013). Predicting premature termination of hospitalised treatment for anorexia nervosa: The roles of therapeutic alliance, motivation, and behaviour change. Eating Behaviors, 14(2), 119–123.

Thaler, L., Israel, M., Antunes, J. M., Sarin, S., Zuroff, D. C., & Steiger, H. (2016). An examination of the role of autonomous versus controlled motivation in predicting inpatient treatment outcome for anorexia nervosa. International Journal of Eating Disorders, 49(6), 626–629.

Tong, J., Dave, N., Mugundu, G. M., Davis, H. W., Gaylinn, B. D., Thorner, M. O. …, & Desai, P. B. (2013). The pharmacokinetics of acyl, desacyl and total ghrelin in healthy human subjects. European Journal of Endocrinology, 168(6), 821–828.

Umeki, S. (1988). Biochemical abnormalities of the serum in anorexia nervosa. The Journal of Nervous and Mental Disease, 176(8), 503–506.

Vandereycken, W. (2003). The place of inpatient care in the treatment of anorexia nervosa: Questions to be answered. International Journal of Eating Disorders, 34(4), 409–422.

Wales, J., Brewin, N., Cashmore, R., Haycraft, E., Baggott, J., Cooper, A., & Arcelus, J. (2016). Predictors of positive treatment outcome in people with anorexia nervosa treated in a specialized inpatient unit: The role of early response to treatment. European Eating Disorders Review, 24(5), 417–424.

Watson, H. J., Yilmaz, Z., Thornton, L. M., Hübel, C., Coleman, J. R., Gaspar, H. A. …, & Medland, S. E. (2019). Genome-wide association study identifies eight risk loci and implicates metabo-psychiatric origins for anorexia nervosa. Nature Genetics, 51(8), 1207–1214.

Weissman, R. S., & Rosselli, F. (2017). Reducing the burden of suffering from eating disorders: Unmet treatment needs, cost of illness, and the quest for cost-effectiveness. Behaviour Research and Therapy, 88, 49–64.

Westmoreland, P., Krantz, M. J., & Mehler, P. S. (2016). Medical complications of anorexia nervosa and bulimia. The American Journal of Medicine, 129(1), 30–37.

Wiseman, C. V., Sunday, S. R., Klapper, F., Harris, W. A., & Halmi, K. A. (2001). Changing patterns of hospitalization in eating disorder patients. International Journal of Eating Disorders, 30(1), 69–74.

